# Translational asymmetry in neuromodulation for substance use disorders: a multi-database bibliometric analysis of primary studies (2000–2025)

**DOI:** 10.64898/2026.08.23.26361148

**Authors:** Samuel Izaias dos Santos Pereira, Matheus Hissa Lourenç Ferreira, Lucas Caseri Câmara, Débora Regina de Aguiar, Rebeca Ferreira de Souza, Tatiana Falcone, Brian S. Barnett, Akhil Anand

## Abstract

**Background:** Substance use disorders (SUDs) remain common worldwide and inadequately treated. Neuromodulation targets neural circuits involved in reward, craving, and cognitive control. However, the primary research literature has not been systematically mapped regarding the relative contributions of clinical and preclinical studies.

**Methods:** We conducted a multi-database bibliometric analysis of primary studies on neuromodulation for SUD. Web of Science, Scopus, and PubMed were searched covering 2000– 2025. After scope classification and exclusion of secondary literature, 810 primary research documents remained. Performance analysis and science mapping were performed with bibliometrix and VOSviewer.

**Results:** Scientific output grew at a compound annual growth rate of 14.16% (2001–2025), accelerating after 2015. Of 810 studies, 82.8% were clinical, 12.5% preclinical, and 4.7% mixed/translational. Alcohol (31.5%) and nicotine/tobacco (25.7%) dominated the literature and were overwhelmingly clinical (>92%), whereas cocaine and opioids retained larger preclinical shares (≈24–27%). Repetitive transcranial magnetic stimulation (rTMS) was the leading modality (31.6%), followed by deep brain stimulation (24.9%) and transcranial direct current stimulation (24.2%). Keyword co-occurrence revealed three clusters: a clinical neuromodulation core, a nicotine/tobacco axis, and a preclinical reward-circuitry module. The United States and China led in output.

**Conclusions:** Neuromodulation research for SUD is expanding rapidly and is heavily skewed toward clinical investigations. A persistent clinical–preclinical asymmetry and limited explicitly translational work constitute structural features of the field. Greater integration between mechanistic and clinical research is needed to advance definitive trials.

## Introduction

Substance use disorders are chronic, relapsing conditions associated with substantial disability, premature mortality, and persistent gaps in treatment access. Recent estimates indicate that more than 53 million people were living with drug use disorders in 2021. Despite this burden, treatment coverage remains limited: data from the United Nations Office on Drugs and Crime indicate that only 1 in 12 individuals with drug use disorders received treatment in 2023, with lower access among women than men. These disparities align with structural limitations in health systems. The World Health Organization’s Mental Health Atlas 2024 reports marked regional differences in specialized workforce capacity, constraining the implementation of device-based interventions across settings [1,2].

Addiction neurobiology provides a circuit-level rationale for neuromodulation. Current models describe addiction as a disorder of distributed cortico-striatal-limbic networks involved in reward, salience, habit formation, interoception, stress regulation, and executive control. Dopamine-mediated signaling within mesolimbic pathways, particularly those involving the nucleus accumbens, enhances the motivational salience of drug-related cues. In parallel, dysfunction within prefrontal control systems impairs regulation of craving, reward processing, and inhibition of compulsive behaviors. The insula and extended amygdala contribute to interoceptive awareness, negative affect, and stress-related relapse vulnerability. Human lesion studies further support this framework: disruption of insular function has been associated with abrupt cessation of cigarette use, indicating a role for interoceptive circuitry as a therapeutic target [3–6].

Neuromodulation acts on neural circuits involved in addiction. Noninvasive techniques, particularly repetitive transcranial magnetic stimulation and transcranial direct current stimulation, have mainly targeted prefrontal regions, especially the dorsolateral prefrontal cortex, to strengthen cognitive control and reduce the influence of drug-related cues. More recent studies have also examined the frontopolar cortex, medial prefrontal cortex, and networks involved in salience processing. Invasive approaches, such as deep brain stimulation, have focused primarily on the nucleus accumbens and the ventral capsule/ventral striatum, which play important roles in reward processing and motivated behavior. The clearest regulatory evidence is in tobacco use disorder, where a deep transcranial magnetic stimulation protocol targeting prefrontal and insular circuitry received U.S. Food and Drug Administration clearance as an aid to short-term smoking cessation following a multicenter randomized trial. Across substance categories, however, reported effect sizes, stimulation targets, and outcome definitions remain heterogeneous [7–12].

Traditional narrative reviews and modality-specific meta-analyses summarize clinical effects but do not define how the primary research literature is organized, how clinical and preclinical domains relate to each other, or which substances and techniques dominate empirical output. Bibliometric science mapping complements these approaches by characterizing the distribution of primary studies, identifying collaboration networks, and quantifying the balance between mechanistic and applied research. These structural patterns inform trial design, funding priorities, and the extent to which findings are likely to generalize across settings [13,14].

Previous bibliometric and mapping studies have examined neuromodulation or addiction-related literatures in isolation, often focusing on a single technique (e.g., rTMS or tDCS), mixing primary and secondary documents, or stopping short of quantifying the balance between clinical and preclinical output. Consequently, the overall structure of the primary literature on neuromodulation for substance use disorders remains unclear, including how mechanistic and clinical research have evolved over time.

This literature can be interpreted using a three-stage translational model: mechanistic plausibility, protocol standardization, and precision deployment. Within this framework, translational maturity reflects progression from a circuit-level rationale to reproducible protocols and, ultimately, to biomarker- or phenotype-informed targeting in defined clinical populations. The central question is not only where the literature concentrates, but whether clinical and preclinical streams have advanced in parallel or remain structurally separated [3–5,7,15–17].

We therefore conducted a multi-database bibliometric analysis restricted to primary studies of neuromodulation in substance use disorders through the end of 2025. Our objectives were to quantify publication growth, map country- and source-level contributions, characterize the thematic structure of the field, and, critically, to measure the clinical–preclinical asymmetry across substances and modalities within a rigorously cleaned corpus of original empirical research.

## Methods

### Study design and reporting framework

We conducted a structured bibliometric analysis combining performance indicators, including productivity, citation dynamics, and growth, with science-mapping methods to examine conceptual and collaboration networks. Reporting followed the Preliminary Guideline for Reporting Bibliometric Reviews of the Biomedical Literature (BIBLIO) [13], with the completed checklist provided in Supplementary Table S1, while the analytical workflow and science-mapping procedures were informed by the Guidelines for Bibliometric-Systematic Literature Reviews (BSLR) [14]. A PRISMA 2020 [18] flow diagram adapted for bibliometric analysis is presented in Supplementary Figure S1.

### Data sources and search strategy

Searches of Web of Science Core Collection, Scopus, and PubMed/MEDLINE were conducted on 3 August 2026, covering the period from 1 January 2000 to 31 December 2025. The strategy combined neuromodulation terms (including “transcranial magnetic stimulation,” rTMS, “transcranial direct current stimulation,” tDCS, “deep brain stimulation,” DBS, theta-burst stimulation, and related techniques) with substance use disorder–related terms (alcohol, nicotine/tobacco, opioids, cocaine, amphetamines/methamphetamine, cannabis, and broader SUD descriptors), using Boolean operators and controlled vocabulary (e.g., MeSH in PubMed) across title, abstract, and keyword fields. Searches were limited to the period 2000–2025. Document-type filters were applied at the search stage to restrict retrieval to original articles (Scopus: DOCTYPE(ar); Web of Science: DT=Article; PubMed: journal article[pt] with exclusion of reviews, meta-analyses, editorials, letters, and related secondary publication types). Full database-specific search strings are provided in Supplementary Table S2.

### Eligibility criteria

Eligible records were primary research documents (original empirical studies) that investigated any form of neuromodulation applied to substance use disorders or substance-related behaviors. Clinical, preclinical, experimental, and translational designs were included. No language restrictions were applied.

Secondary literature was excluded: systematic reviews, meta-analyses, narrative reviews, editorials, letters, commentaries, conference abstracts, protocols, and guidelines. Records that did not address both neuromodulation and substance-related outcomes were classified as out of scope and removed. Document type and study design (clinical, preclinical, or mixed/translational) were retained as analytic variables.

### Data processing and cleaning

Records were exported from each database, merged, and deduplicated using DOI matching, normalized title–year comparison, and targeted manual review of residual ambiguous cases. After initial filters (publication year, document type, and retraction status), unique records underwent automated and manual scope classification. Studies retained after scope screening were further inspected to remove residual reviews and near-duplicate records. The final analytic corpus comprised 810 primary research documents.

Metadata were standardized for publication year, source, authorship, affiliations, and keywords. Studies were classified as clinical, preclinical, or mixed/translational using title and abstract (and full text when necessary). Records were labeled clinical when they reported human participants, patients, or clinical samples; preclinical when they were restricted to animal models, ex vivo preparations, or purely mechanistic non-human designs; and mixed/translational when both human and non-human evidence were integrated within the same primary report, or when the study explicitly bridged mechanistic and clinical endpoints. Ambiguous cases were resolved by manual adjudication. Substance categories and neuromodulation modalities were assigned through controlled text matching and were allowed to be non-exclusive (a single document could contribute to more than one category).

### Statistical analysis

Analyses were conducted in R version 4.5.1 [19] (bibliometrix package [20]), with additional visualization in ggplot2 [21] and network mapping in VOSviewer [22]. Annual production trends were summarized with a linear trend and compound annual growth rate (CAGR) calculated from the first year with recorded output (2001) to 2025. Performance indicators included most productive authors, countries, and sources, as well as citation metrics.

Keyword co-occurrence networks were built from cleaned author keywords after removal of generic demographic and methodological terms and residual comorbidity labels unrelated to the core SUD focus. Association-strength normalization and Louvain clustering were applied. Country collaboration networks were derived from co-authorship and affiliation data. Substance- and modality-specific volumes and clinical–preclinical shares were computed from the classified corpus.

### Methodological quality assessment

Risk-of-bias assessment of individual studies was not performed because the objective was structural mapping of the primary research landscape rather than evidence synthesis of clinical efficacy. Findings should be interpreted as indicators of research activity, conceptual organization, translational balance, and collaboration patterns.

## Results

### Search results and corpus characteristics

The multi-database search (Web of Science, Scopus and PubMed) followed by automated and manual cleaning yielded a final analytical corpus of 810 primary research documents published between 2000 and 2025. After removal of duplicates, out-of-scope records, retracted papers and all forms of secondary literature (systematic reviews, meta-analyses, narrative reviews and editorials), the corpus contained exclusively original empirical studies. Of these, 671 (82.8%) were classified as clinical, 101 (12.5%) as preclinical, and 38 (4.7%) as mixed or translational.

Scientific output grew at a CAGR of 14.16% from 2001 to 2025. Annual production remained modest until approximately 2012 (fewer than 20 documents per year), accelerated thereafter, and reached 97 publications in 2025 (Figure 1). The average number of citations per document was 33.4, and 30.9% of documents involved international co-authorship.

**Figure 1.**
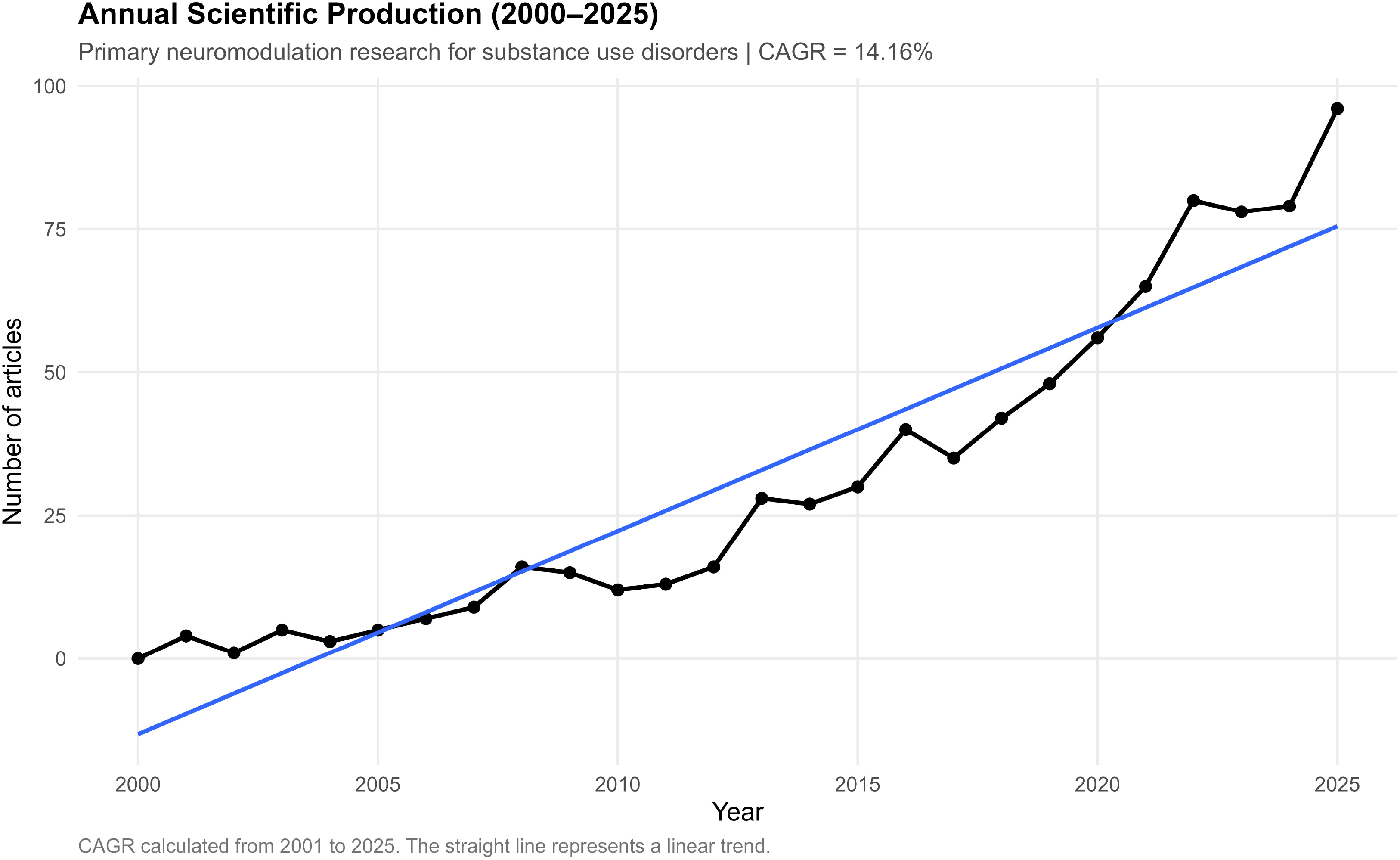
Annual scientific production of primary studies on neuromodulation for substance use disorders (2001–2025) Annual number of primary research documents (solid line with points). The additional solid line represents the linear trend. Compound annual growth rate (CAGR) = 14.16%, calculated from 2001 (first year with output) to 2025. N = 810.

### Most productive authors, countries and sources

The five most productive authors were Li X (25 documents), George MS (20), Hanlon CA (20), Fregni F (18) and Zangen A (18) (Table 1).

**Table 1.** Most productive authors in neuromodulation research for substance use disorders.

| Rank | Author | Documents | Fractionalized documents |
| --- | --- | --- | --- |
| 1 | Li X | 25 | 3.24 |
| 2 | George MS | 20 | 3.45 |
| 3 | Hanlon CA | 20 | 3.41 |
| 4 | Fregni F | 18 | 2.67 |
| 5 | Zangen A | 18 | 2.19 |
| 6 | Wang X | 17 | — |
| 7 | Daskalakis ZJ | 16 | 2.18 |
| 8 | Blumberger DM | 14 | — |
| 9 | Zhang J | 14 | — |
| 10 | Li Y | 13 | — |

**Note:** Top 10 authors by number of primary studies (2001–2025). Fractionalized counts are reported when available from the bibliometric output. N = 810.

The geographic distribution of corresponding authors was led by the United States (186 documents, 25.7%), followed by China (115, 15.9%), Germany (56), Iran (53) and Italy (49) (Table 2). Brazil ranked eighth (23 documents) yet exhibited the highest multiple-country publication ratio among the top ten countries (MCP = 0.783), reflecting a strongly collaborative national profile. Canada and Israel recorded the highest average citations per article (89.4 and 167.0, respectively). The country collaboration network showed a core centered on the United States, with strong links to China, Canada, European countries and Brazil (Figure 2). The global distribution of scientific production is shown in Supplementary Figure S2.

**Figure 2.**
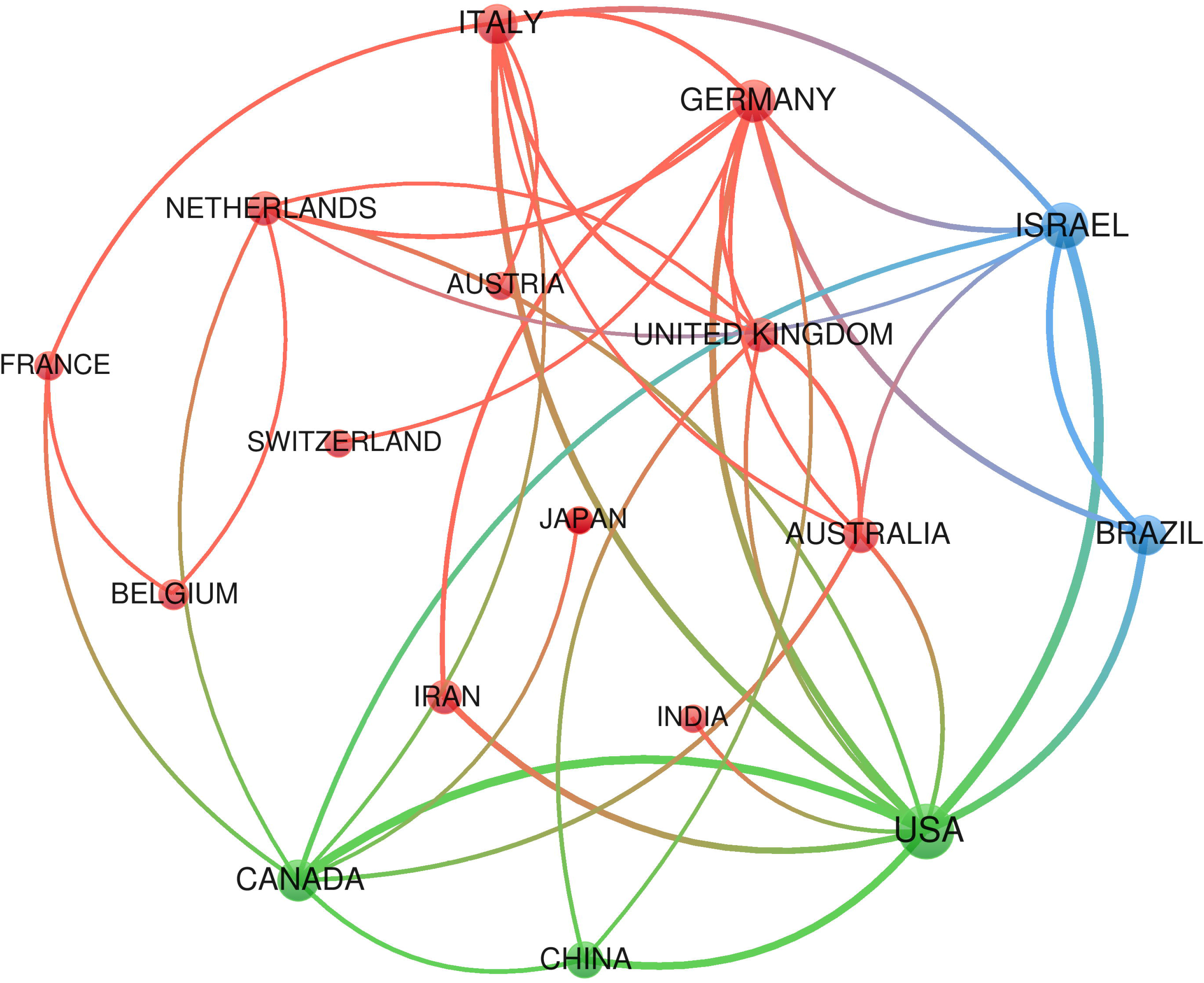
Country collaboration network in neuromodulation research for substance use disorders Co-authorship network of the leading countries. Node size reflects publication volume; link thickness reflects collaboration intensity. N = 810.

**Table 2.** Leading countries by scientific production and international collaboration.

| **Rank** | **Country** | **Documents** | **%** | **SCP** | **MCP** | **MCP ratio** |
| --- | --- | --- | --- | --- | --- | --- |
| 1 | USA | 186 | 25.7 | 146 | 40 | 0.215 |
| 2 | China | 115 | 15.9 | 91 | 24 | 0.209 |
| 3 | Germany | 56 | 7.7 | 37 | 19 | 0.339 |
| 4 | Iran | 53 | 7.3 | 42 | 11 | 0.208 |
| 5 | Italy | 49 | 6.8 | 27 | 22 | 0.449 |
| 6 | Canada | 39 | 5.4 | 19 | 20 | 0.513 |
| 7 | Netherlands | 26 | 3.6 | 18 | 8 | 0.308 |
| 8 | Brazil | 23 | 3.2 | 5 | 18 | 0.783 |
| 9 | France | 22 | 3.0 | 13 | 9 | 0.409 |
| 10 | Belgium | 19 | 2.6 | 13 | 6 | 0.316 |

**Abbreviations:** MCP, multiple-country publications; SCP, single-country publications. The
MCP ratio represents the proportion of publications involving international collaboration.

The journals that published the largest number of primary studies were Frontiers in Psychiatry (38), Drug and Alcohol Dependence (30), Brain Stimulation (29), Addiction Biology (21) and Neuropsychopharmacology (18) (Table 3). This distribution indicates that the field is shared between specialized addiction outlets and journals focused on neuromodulation and biological psychiatry. The cumulative trajectory of the leading journals is shown in Supplementary Figure S3.

**Table 3.** Journals with the highest number of publications.

| **Rank** | **Journal** | **Documents** |
| --- | --- | --- |
| 1 | *Frontiers in Psychiatry* | 38 |
| 2 | *Drug and Alcohol Dependence* | 30 |
| 3 | *Brain Stimulation* | 29 |
| 4 | *Addiction Biology* | 21 |
| 5 | *Neuropsychopharmacology* | 18 |
| 6 | *PLOS ONE* | 13 |
| 7 | *Scientific Reports* | 13 |
| 8 | *Biological Psychiatry* | 11 |
| 9 | *Frontiers in Human Neuroscience* | 11 |
| 10 | *Journal of Psychiatric Research* | 10 |

### Distribution by substance category

Alcohol-related research constituted the largest category (255 documents, 31.5% of the corpus), followed by nicotine/tobacco (208, 25.7%), opioids (141, 17.4%), cocaine (122, 15.1%), other stimulants (122, 15.1%) and cannabis (60, 7.4%) (Figure 3). Categories are not mutually exclusive; individual studies frequently addressed more than one substance. Temporal trends showed sustained growth for alcohol and nicotine across the entire period, with a sharper rise after 2015, while cannabis remained comparatively marginal until the most recent years (Supplementary Figure S4).

**Figure 3.**
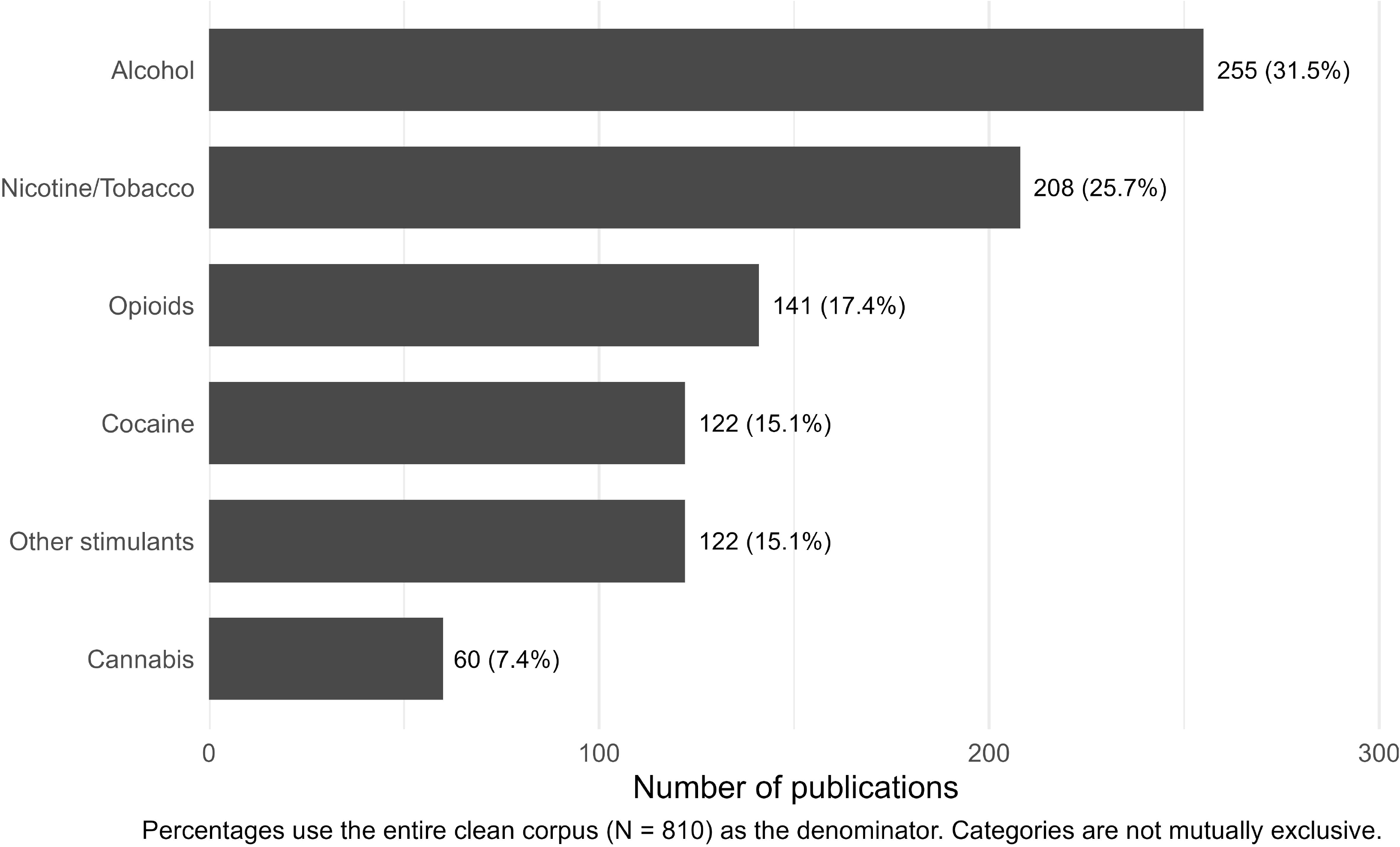
Publication volume by substance category in neuromodulation research for substance use disorders Number and percentage of primary studies addressing each substance category. Percentages use the full corpus (N = 810) as denominator. Categories are not mutually exclusive.

### Clinical–preclinical asymmetry across substances

A marked translational imbalance characterized the corpus. After exclusion of mixed/translational studies, clinical investigations accounted for more than 90% of the literature on alcohol (92.7%) and nicotine/tobacco (95.5%). In contrast, cocaine and opioids retained substantially larger preclinical shares (26.7% and 23.7%, respectively) (Figure 4). Other stimulants occupied an intermediate position (clinical share 84.0%). Only 38 documents (4.7% of the full corpus) were classified as mixed or explicitly translational, indicating limited integration within individual primary reports.

**Figure 4.**
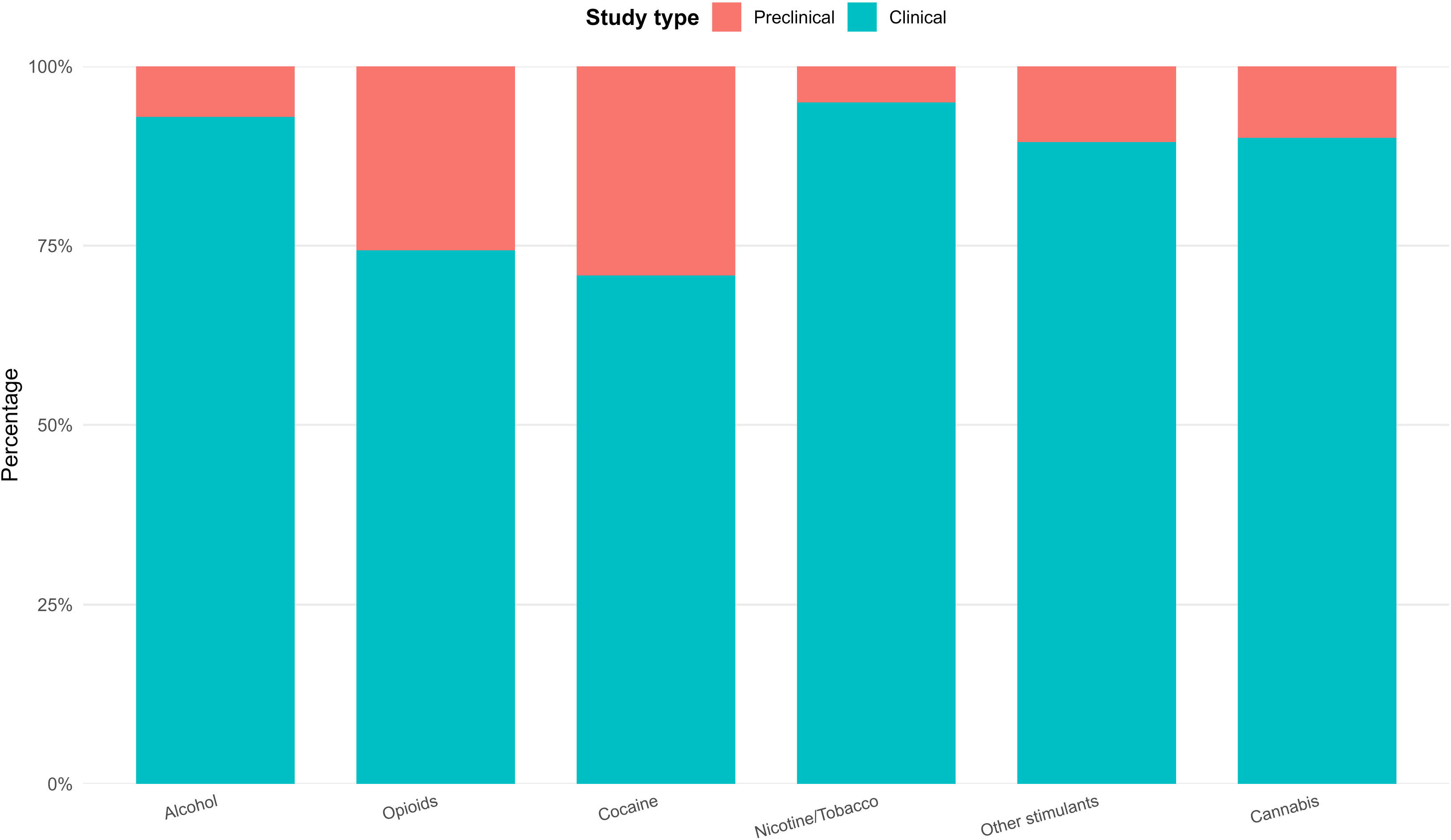
Clinical versus preclinical research share by substance category Proportion of clinical and preclinical primary studies within each substance category, calculated after excluding mixed/translational records from the denominator of that substance. N = 810 for the full corpus; denominators vary by substance.

### Neuromodulation modalities

When both broad and specific descriptors were considered, transcranial magnetic stimulation (TMS) in any form appeared in 50.5% of documents. More narrowly defined repetitive TMS (rTMS) was identified in 31.6%, deep brain stimulation (DBS) in 24.9%, and transcranial direct current stimulation (tDCS) in 24.2%. Theta-burst stimulation (TBS) already represented 8.1% of the corpus. Because rTMS and TBS are nested within the broader TMS literature, these categories are not mutually exclusive; focused ultrasound and other emerging techniques remained infrequent (<3%). Temporal trends for the full set of modalities, including less frequent techniques, are presented in Supplementary Figure S5.

Temporal trajectories of the five principal modalities revealed a clear inflection for rTMS after 2018–2019, after which it became the dominant technique by volume (Figure 5). DBS and tDCS displayed steadier, approximately linear growth throughout the study period. TBS, although still modest in absolute numbers, exhibited a steep recent increase consistent with growing methodological interest in shorter, high-frequency protocols.

**Figure 5.**
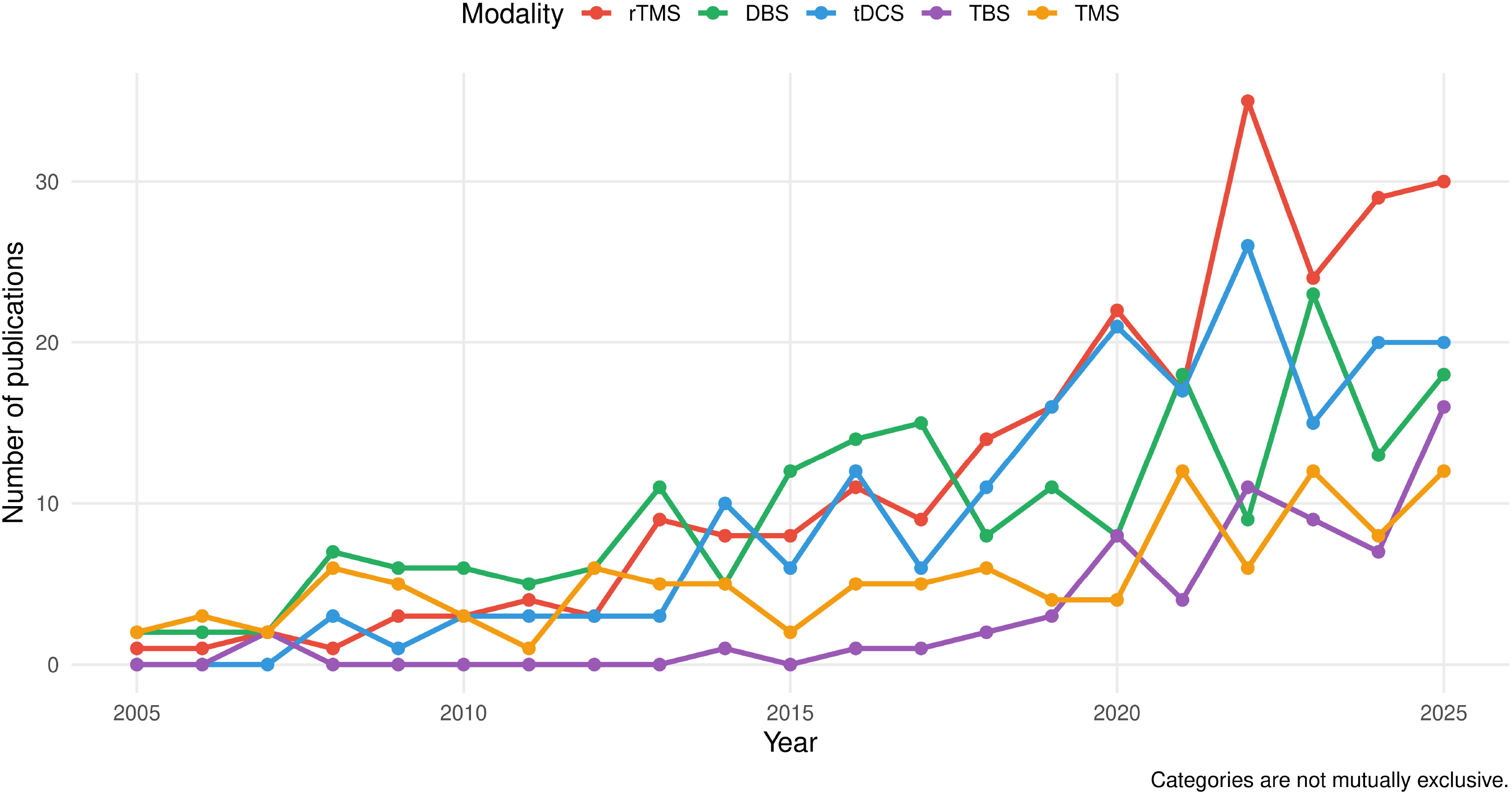
Temporal trends in publications by main neuromodulation modalities. Annual number of primary studies for rTMS, DBS, tDCS, TBS, and TMS (2005–2025). Categories are not mutually exclusive. N = 810.

### Conceptual structure of the field

Author-keyword co-occurrence analysis of the cleaned corpus produced three coherent clusters (Figure 6). The first cluster centered on clinical neuromodulation and comprised terms such as transcranial magnetic stimulation, repetitive transcranial magnetic stimulation, transcranial direct current stimulation, craving, dorsolateral prefrontal cortex and prefrontal cortex. The second cluster was spatially and thematically distinct and corresponded to the nicotine/tobacco literature (smoking, nicotine, tobacco and tobacco use disorder). The third cluster linked reward-related and substance-specific terms with a more preclinical orientation and included nucleus accumbens, dopamine, morphine, cocaine, addiction, relapse, methamphetamine and alcohol. The relative isolation of the nicotine/tobacco axis and the peripheral position of intermittent theta-burst stimulation were notable structural features of the network. The temporal peak of key author keywords further illustrated the recent concentration of clinical neuromodulation terms (Supplementary Figure S6).

**Figure 6.**
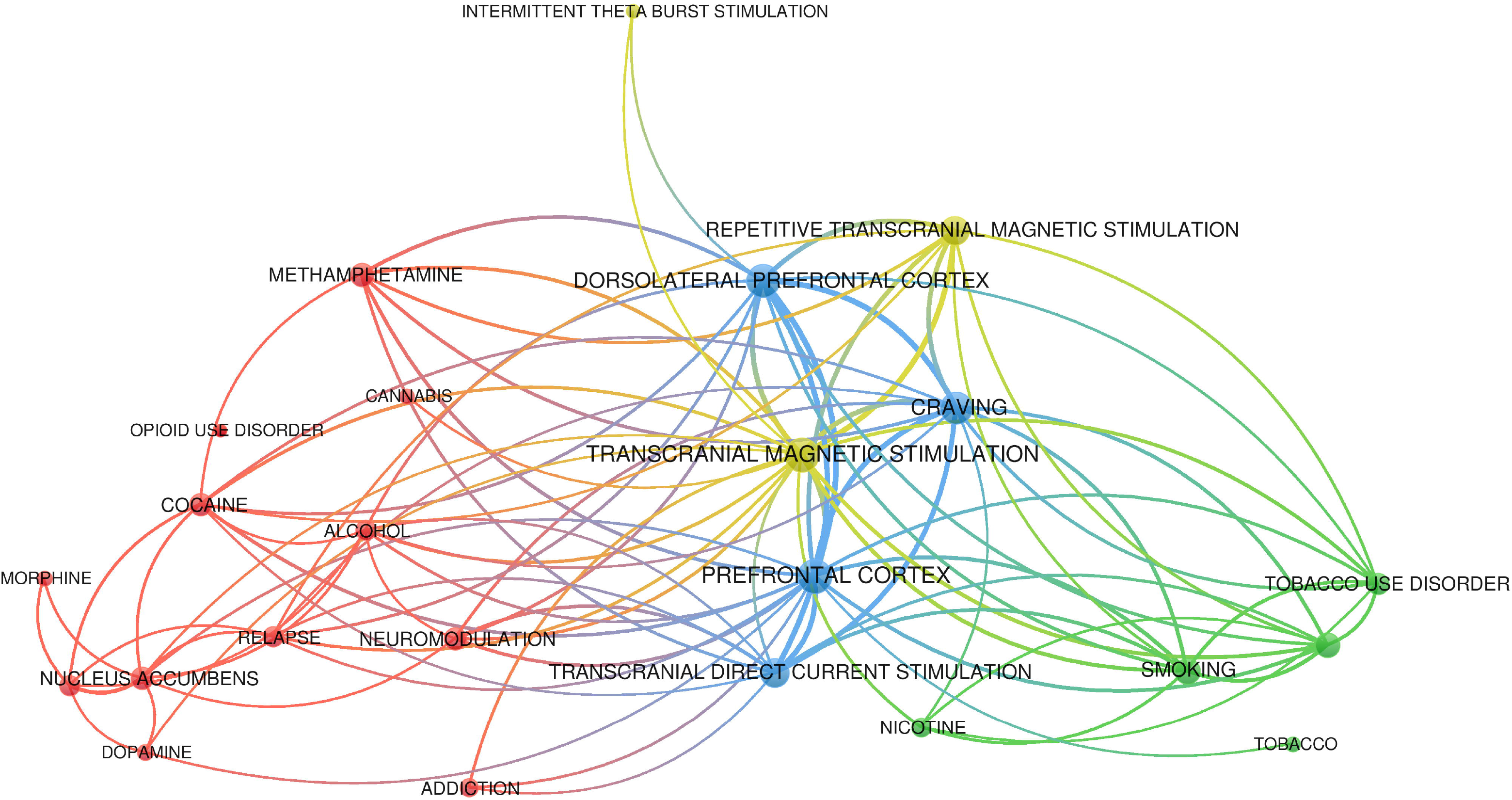
Author-keyword co-occurrence network of primary studies on neuromodulation for substance use disorders. Network of the top cleaned author keywords (association-strength normalization). Node size reflects occurrence frequency; colors indicate clusters. Three main clusters are visible: clinical neuromodulation (center), nicotine/tobacco (right), and reward/substance-related terms (left). N = 810.

## Discussion

### Principal findings

This bibliometric analysis of 810 primary studies provides the most rigorously cleaned map to date of neuromodulation research applied to substance use disorders. Three findings stand out. First, the field has grown rapidly and continuously (CAGR ≈ 14%), with acceleration after 2015 that coincides with broader clinical adoption of non-invasive brain stimulation [7]. Second, research activity is heavily skewed toward clinical investigations, especially for alcohol and nicotine, while cocaine and opioid research retain a more balanced preclinical component. Third, rTMS has emerged as the dominant modality in recent years, surpassing both DBS and tDCS in annual output.

### Translational asymmetry as a structural feature of the field

The most important finding is the marked imbalance between clinical and preclinical research. More than four-fifths of the primary studies are clinical, although this proportion varies by substance. For alcohol and nicotine, which already have well-established clinical trial infrastructures and regulatory pathways, clinical studies account for more than 92% of the literature [8,9]. In contrast, cocaine and opioids, whose clinical development faces greater logistical and ethical constraints, continue to draw a substantial fraction of their evidence from animal models [10,11]. The scarcity of explicitly mixed or translational designs (only 4.7% of the corpus) suggests limited integration within individual primary reports. This metric does not capture translation that may occur across separate papers within the same research program. Even so, the marked imbalance in study type across substances indicates that clinical and mechanistic streams have not developed symmetrically. Funding mechanisms and collaborative platforms that deliberately incentivize bidirectional work may help reduce this structural gap.

### Modality dynamics and emerging techniques

The rapid rise of rTMS after 2018 is consistent with accumulating clinical evidence, improved coil designs, and the relative practicality of the technique compared with invasive approaches [7,8]. The concurrent growth of theta-burst protocols suggests that the field is already moving toward more time-efficient stimulation paradigms. DBS remains an important modality in absolute volume, particularly in studies involving reward-related targets, but its clinical uptake in addiction is constrained by invasiveness and patient selection. tDCS occupies an intermediate position: accessible and widely studied, yet still seeking definitive large-scale efficacy data. Focused ultrasound and related emerging modalities are present but still peripheral, indicating that the next wave of technological innovation has not yet translated into substantial primary-study volume.

### Geographic and collaborative patterns

Scientific production is concentrated in the United States and China, which together account for more than 40% of corresponding-author output. Nevertheless, several mid-sized research systems display high rates of international collaboration. Brazil’s MCP ratio of 0.78 is the highest among the top ten countries, suggesting that Brazilian groups are already embedded in global networks despite lower absolute volume. Canada and Israel achieve disproportionately high citation impact, pointing to specialized, high-quality niches. These patterns imply that capacity-building strategies should prioritize the strengthening of existing collaborative links rather than solely the expansion of domestic output.

### Conceptual organization of knowledge

The three-cluster structure recovered from author keywords partially mirrors the translational divide observed in study-type classification. A clinical neuromodulation core (centered on TMS, tDCS, craving and prefrontal targets) is distinguishable from a cluster of reward- and substance-related terms (including nucleus accumbens, dopamine, cocaine, morphine, addiction and relapse), while the nicotine/tobacco literature forms a relatively self-contained module. This modularity may facilitate specialized progress within each domain, yet it also risks fragmentation of the broader knowledge base. Intermittent theta-burst stimulation remains peripheral in the network, consistent with its more recent entry into the addiction literature and its still-limited co-occurrence with established clinical and mechanistic terms.

### Strengths and limitations

Major strengths of the present work include the restriction to primary studies, the transparent multi-stage cleaning pipeline, the explicit quantification of clinical–preclinical balance across substances, and the use of both performance and science-mapping indicators. Limitations must also be acknowledged. Keyword and modality assignments are non-exclusive; percentages therefore over-count multi-label documents when summed across categories. Author-keyword coverage was incomplete (approximately 18% of records lacked author keywords), although Keywords Plus partially compensated. Country assignment relied on corresponding-author and affiliation data and may under-represent multi-national teams. Finally, the analysis maps research activity rather than clinical efficacy; high publication volume should not be interpreted as evidence of therapeutic superiority.

### Implications and future directions

The findings carry three practical implications. First, funding agencies and journal editors should recognize the translational asymmetry as a structural feature of the field and create incentives for studies that explicitly link mechanistic and clinical endpoints. Second, the rapid consolidation of rTMS as the leading modality warrants focused evidence synthesis and, where appropriate, comparative trials against tDCS and TBS [12,15]. Third, the relatively self-contained nicotine literature and the still-modest cannabis output identify opportunities for cross-substance learning and for expanding primary research on less-studied substances. Future bibliometric updates will be needed to track whether the recent acceleration of TBS and the slow emergence of focused ultrasound alter the current hierarchy of modalities.

## Conclusions

Neuromodulation research on substance use disorders has expanded rapidly over the past two decades and is now dominated by clinical investigations, particularly of alcohol and nicotine, and by repetitive transcranial magnetic stimulation. A persistent clinical–preclinical asymmetry, the limited volume of explicitly translational work, and the modular organization of the knowledge base constitute the central structural features of the field. Addressing these features through deliberate bridging strategies may help ensure that the growing body of primary evidence translates into improved treatments for substance use disorders.

## Data Availability

All data produced in the present study are available upon reasonable request to the authors.

## Declarations

### Ethics Approval and Consent to Participate

Not applicable.

### Consent for Publication

Not applicable.

### Availability of Data and Materials

The datasets generated and/or analyzed during the current study are available from the corresponding author on reasonable request.

### Authors’ Contributions

SISP: Conceptualization, Methodology, Writing – original draft, Writing – review & editing.

MHLF: Methodology, Software, Formal analysis, Data curation, Visualization, Writing – original draft, Writing – review & editing.

LCC: Conceptualization, Methodology, Writing – original draft. DRA: Writing – original draft.

RFS: Writing – original draft.

TF: Writing – original draft, Writing – review & editing.

BSB: Writing – original draft, Writing – review & editing, Supervision. AA: Writing – original draft, Writing – review & editing, Supervision. All authors read and approved the final manuscript.

## Acknowledgements

None.

