## Supplementary Material for "Translational asymmetry in neuromodulation for substance use disorders: a multi-database bibliometric analysis of primary studies (2000–2025)"

**Supplementary Table S1.** The BIBLIO Checklist for Reporting Bibliometric Reviews of the Biomedical Literature

| Section/Topic | Item No. | Checklist item | Reported on page No. |
|---|---|---|---|
| **Title** | | | |
| Identification | 1 | Identify the report as a bibliometric review in the title. | 1 |
| Issues/topics | 2 | Indicate the key issues/topics under investigation and coverage of time period. | 1 (título: 2000–2025); também 1–3 |
| **Abstract** | | | |
| Structured summary | 3 | Structured summary including, as applicable: background, methods, results (key findings), and conclusions. | 1 |
| **Introduction/Background** | | | |
| Justification/Rationale/Explanation | 4 | Present review of existing knowledge and epidemiological information. | 1–3 |
| Objectives | 5 | Statement of the objective(s) or question(s). | 3 |
| **Methods** | | | |
| Search engines (data sources) | 6 | Describe all information sources, such as electronic databases, contact with study authors, trial registers, or other grey literature sources. | 3–4 |
| Search strategy | 7 | Keywords and systematization criteria—date of search, language, and type of document—for the search. | 3–4 |
| Time period | 8 | The period that the review covers and the justification. | 1, 3–4 |
| Eligibility criteria | 9 | Describe all inclusion and exclusion criteria; languages; study design, type of publication, and time period. | 4 |
| Data refinement (data selection procedure) | 10 | Remove irrelevant articles; inspection to eliminate duplicate | 4 |

| Section/Topic | Item No. | Checklist item | Reported on page No. |
|---|---|---|---|
| | | and unrelated articles after evaluation of the title, abstract, and content. | |
| Quality assessment (optional) | 11 | Assessment of papers by three authors and the use of assessing checklists. | 5 (seção *Methodological quality assessment*: RoB não realizado, com justificativa) |
| Data synthesis | 12 | Describe the methods used for summarizing, handling, synthesis, tabulations, or schematic displays. Describe how the data were analysed. | 4–5 |
| **Results** | | | |
| Descriptive findings (statistics) | 13 | Provide details of the search and selection process in a flow diagram. Report the number of citations retrieved, including number of publications, year of publication, type of documents, country of publication, articles with the highest impact, most impactful authors, most impactful articles, authors with the highest production, top journals, top institutions, etc. | 5–6 (fluxo: Suppl. Fig. S1, citado na p. 3) |
| Schematic map and trend | 14 | Summarize and/or present schematic maps and trends using appropriate software to present citations, journals, authors, top journals, time trends, emerging literature, and any relevant indicators, as applicable. | 5–7 (Figs. 1–6; legendas na p. 14) |
| Tabulation and summarizing the findings | 15 | Studies under consideration could be summarized and organized by different subtitles and different scenarios. Results need to be presented in separate tables covering each subtitle. | 5–6 (Tables 1–3: autores, países, periódicos) |
| Synthesis of findings | 16 | Synthesize the findings as much as possible, find the gap, and | 8–10 |

| Section/Topic | Item No. | Checklist item | Reported on page No. |
|---|---|---|---|
| | | propose a model, hypothesis, etc., if applicable. | |
| **Discussion** | | | |
| Summary of evidence | 17 | Summarize the main findings. The findings should be presented in more “general” or “accessible” terms. | 8 |
| Interpretation | 18 | Include interpretation consistent with the results. Explanations for observed outcomes, similarities, and differences reported would be essential. | 8–10 |
| Strengths and limitations | 19 | Discuss the strengths and limitations. | 9–10 |
| Conclusion(s) | 20 | Provide a general interpretation of the results with respect to the review questions and objectives, as well as potential implications. | 10 |

Rights and permissions: The original source of the checklist is: Montazeri A, Mohammadi S, M.Hesari P, Ghaemi M, Riazi H, Sheikhi-Mobarakeh Z. Preliminary guideline for reporting bibliometric reviews of the biomedical literature (BIBLIO): a minimum requirements. *Systematic Reviews* 2023; 12: 239. doi.org/10.1186/s13643-023-02410-2 The article is licensed under a Creative Commons Attribution 4.0 International License (http://creativecommons.org/licenses/by/4.0/). A changes was made to the original checklist to add in full references to the cited sources.

**Supplementary Table S2.** Search strategies used in Web of Science, Scopus, and PubMed. Searches were conducted on 3 August 2026 and covered literature published from 2000 to 31 December 2025.

| Database | Search strategy |
|---|---|
| **Web of Science** | TS=( "transcranial magnetic stimulation" OR "repetitive transcranial magnetic stimulation" OR rTMS OR "deep transcranial magnetic stimulation" OR dTMS OR "theta burst stimulation" OR "intermittent theta burst stimulation" OR "continuous theta burst stimulation" OR iTBS OR cTBS OR "transcranial direct current stimulation" OR tDCS OR "transcranial alternating current stimulation" OR tACS OR "transcranial random noise stimulation" OR tRNS OR "transcranial electrical stimulation" OR "transcranial electric stimulation" OR "deep brain stimulation" OR DBS OR "transcranial focused ultrasound" OR "focused ultrasound neuromodulation" OR "low-intensity focused ultrasound" OR "low intensity focused ultrasound" OR "transcranial ultrasound stimulation" OR "temporal interference stimulation" OR neuromodulat*)ANDTS=( "substance use disorder*" OR "substance-related disorder*" OR "drug use disorder*" OR "addictive disorder*" OR "drug addiction" OR "drug dependence" OR "substance addiction" OR "substance dependence" OR "substance abuse" OR polysubstance OR "polysubstance use" OR "polydrug use" OR ( (alcohol* OR ethanol OR tobacco OR nicotine OR smok* OR opioid* OR opiate* OR heroin OR morphine OR fentanyl OR cocaine OR crack OR cannabis OR marijuana OR marihuana OR hashish OR "synthetic cannabinoid*" OR amphetamine* OR methamphetamine* OR stimulant* OR benzodiazepine* OR sedative* OR hypnotic* OR anxiolytic* OR hallucinogen* OR phencyclidine OR PCP OR MDMA OR ecstasy OR ketamine OR inhalant* OR solvent*) AND (addict* OR dependenc* OR abuse OR misuse OR "use disorder*" OR craving OR withdrawal OR relapse OR abstinen* OR cessation) ))AND PY=2000-2025AND DT=(Article) |
| **Scopus** | TITLE-ABS-KEY( "transcranial magnetic stimulation" OR "repetitive transcranial magnetic stimulation" OR rTMS OR "deep transcranial magnetic stimulation" OR dTMS OR "theta burst stimulation" OR "intermittent theta burst stimulation" OR "continuous theta burst stimulation" OR iTBS OR cTBS OR "transcranial direct current stimulation" OR tDCS OR "transcranial alternating current stimulation" OR tACS OR "transcranial random noise stimulation" OR tRNS OR "transcranial electrical stimulation" OR "transcranial electric stimulation" OR "deep brain stimulation" OR DBS OR "transcranial focused ultrasound" OR "focused ultrasound neuromodulation" OR "low-intensity focused ultrasound" OR "low intensity focused ultrasound" OR "transcranial ultrasound stimulation" OR "temporal interference stimulation" OR neuromodulat*)ANDTITLE-ABS-KEY( "substance use disorder*" OR "substance-related disorder*" OR "drug use disorder*" OR "addictive disorder*" OR "drug addiction" OR "drug dependence" OR "substance addiction" OR "substance dependence" OR "substance abuse" OR polysubstance OR "polysubstance use" OR "polydrug use" OR ( (alcohol* OR ethanol OR tobacco OR nicotine OR smok* OR opioid* OR opiate* OR heroin OR morphine OR fentanyl OR cocaine OR crack OR cannabis OR marijuana OR marihuana OR hashish OR |

| Database | Search strategy |
| --- | --- |
| | "synthetic cannabinoid*" OR amphetamine* OR methamphetamine* OR stimulant* OR benzodiazepine* OR sedative* OR hypnotic* OR anxiolytic* OR hallucinogen* OR phencyclidine OR PCP OR MDMA OR ecstasy OR ketamine OR inhalant* OR solvent*) AND (addict* OR dependenc* OR abuse OR misuse OR "use disorder*" OR craving OR withdrawal OR relapse OR abstinen* OR cessation) ))AND PUBYEAR > 1999 AND PUBYEAR < 2026AND DOCTYPE(ar) |
| **PubMed** | ( "Transcranial Magnetic Stimulation"[Mesh] OR "Transcranial Direct Current Stimulation"[Mesh] OR "Deep Brain Stimulation"[Mesh] OR "transcranial magnetic stimulation"[tiab] OR "repetitive transcranial magnetic stimulation"[tiab] OR rTMS[tiab] OR "deep transcranial magnetic stimulation"[tiab] OR dTMS[tiab] OR "theta burst stimulation"[tiab] OR "intermittent theta burst stimulation"[tiab] OR "continuous theta burst stimulation"[tiab] OR iTBS[tiab] OR cTBS[tiab] OR "transcranial direct current stimulation"[tiab] OR tDCS[tiab] OR "transcranial alternating current stimulation"[tiab] OR tACS[tiab] OR "transcranial random noise stimulation"[tiab] OR tRNS[tiab] OR "transcranial electrical stimulation"[tiab] OR "transcranial electric stimulation"[tiab] OR "deep brain stimulation"[tiab] OR DBS[tiab] OR "transcranial focused ultrasound"[tiab] OR "focused ultrasound neuromodulation"[tiab] OR "low-intensity focused ultrasound"[tiab] OR "low intensity focused ultrasound"[tiab] OR "transcranial ultrasound stimulation"[tiab] OR "temporal interference stimulation"[tiab] OR neuromodulat*[tiab] ) AND ( "Substance-Related Disorders"[Mesh] OR "Alcohol-Related Disorders"[Mesh] OR "Alcoholism"[Mesh] OR "Tobacco Use Disorder"[Mesh] OR "Smoking Cessation"[Mesh] OR "Opioid-Related Disorders"[Mesh] OR "Cocaine-Related Disorders"[Mesh] OR "Amphetamine-Related Disorders"[Mesh] OR "Marijuana Abuse"[Mesh] OR "Inhalant Abuse"[Mesh] OR "Phencyclidine Abuse"[Mesh] OR "substance use disorder"[tiab] OR "substance use disorders"[tiab] OR "substance-related disorder"[tiab] OR "substance-related disorders"[tiab] OR "drug use disorder"[tiab] OR "drug use disorders"[tiab] OR "addictive disorder"[tiab] OR "addictive disorders"[tiab] OR "drug addiction"[tiab] OR "drug dependence"[tiab] OR "substance addiction"[tiab] OR "substance dependence"[tiab] OR "substance abuse"[tiab] OR polysubstance[tiab] OR "polysubstance use"[tiab] OR "polydrug use"[tiab] OR ( (alcohol*[tiab] OR ethanol[tiab] OR tobacco[tiab] OR nicotine[tiab] OR smok*[tiab] OR opioid*[tiab] OR opiate*[tiab] OR heroin[tiab] OR morphine[tiab] OR fentanyl[tiab] OR cocaine[tiab] OR crack[tiab] OR cannabis[tiab] OR marijuana[tiab] OR marihuana[tiab] OR hashish[tiab] OR "synthetic cannabinoid"[tiab] OR "synthetic cannabinoids"[tiab] OR amphetamine*[tiab] OR methamphetamine*[tiab] OR stimulant*[tiab] OR benzodiazepine*[tiab] OR sedative*[tiab] OR hypnotic*[tiab] OR anxiolytic*[tiab] OR hallucinogen*[tiab] OR phencyclidine[tiab] OR PCP[tiab] OR MDMA[tiab] OR ecstasy[tiab] OR ketamine[tiab] OR inhalant*[tiab] OR solvent*[tiab]) AND (addict*[tiab] OR dependenc*[tiab] OR abuse[tiab] OR misuse[tiab] OR "use disorder"[tiab] OR "use disorders"[tiab] OR craving[tiab] OR withdrawal[tiab] OR relapse[tiab] OR abstinen*[tiab] OR cessation[tiab]) ) ) AND ("2000/01/01"[Date - Publication] : "2025/12/31"[Date - Publication]) AND (journal article[pt]) NOT (review[pt] OR systematic review[pt] OR meta-analysis[pt] OR editorial[pt] OR |

| Database | Search strategy |
|---|---|
| | letter[pt] OR comment[pt] OR guideline[pt] OR practice guideline[pt] OR news[pt] OR published erratum[pt] OR retraction notice[pt] OR "clinical trial protocol"[pt]) |

**Supplementary Figure S1.** PRISMA 2020 flow diagram of study identification and selection Records identified from Web of Science, Scopus and PubMed, followed by deduplication, scope screening, exclusion of secondary literature and residual cleaning. Final analytic corpus: 810.

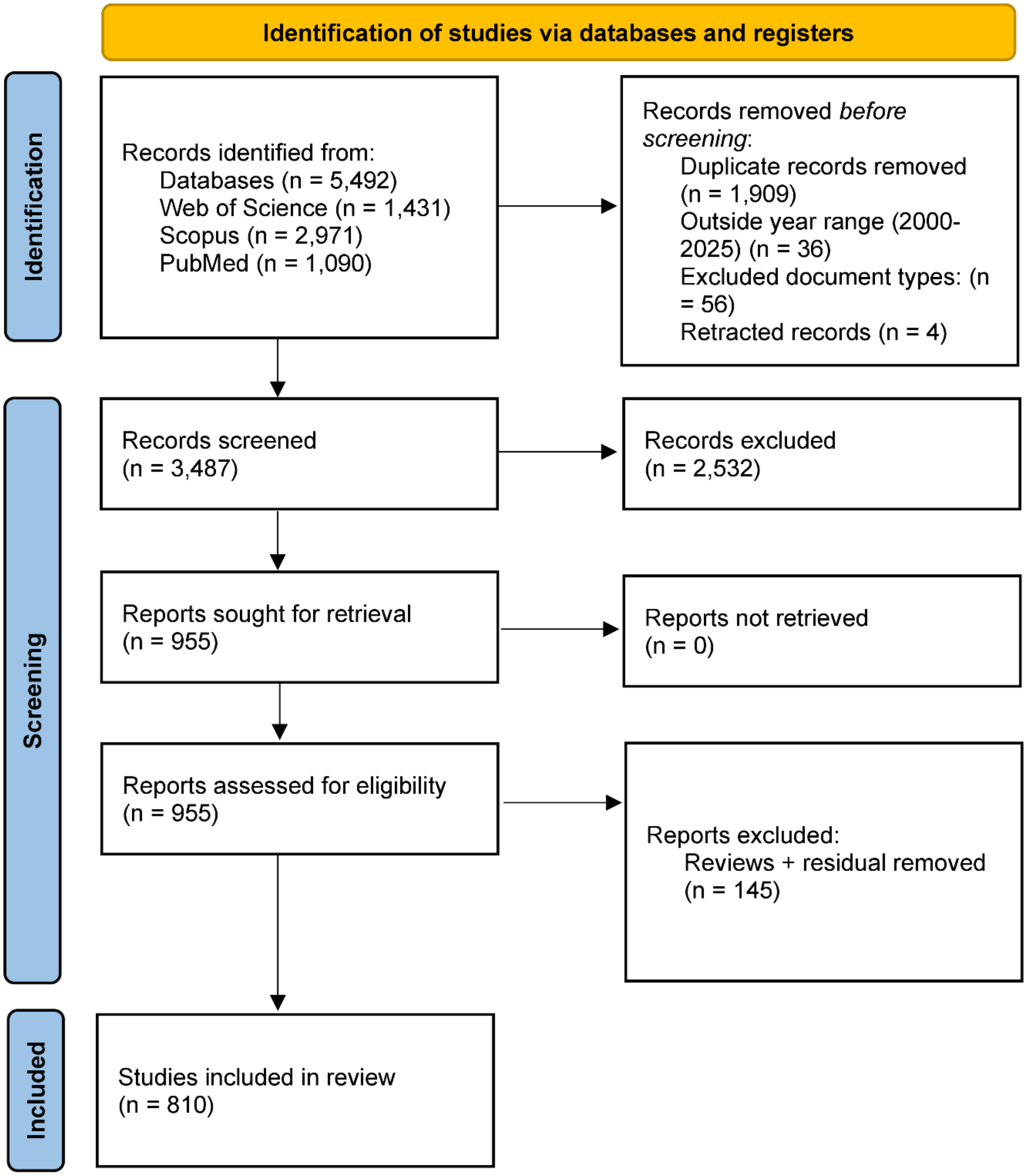

**Supplementary Figure S2.** Global distribution of scientific production.

Country-level publication counts based on all available author affiliations. Each document is counted once per country. Hong Kong was merged with China for polygon mapping only. N = 810.

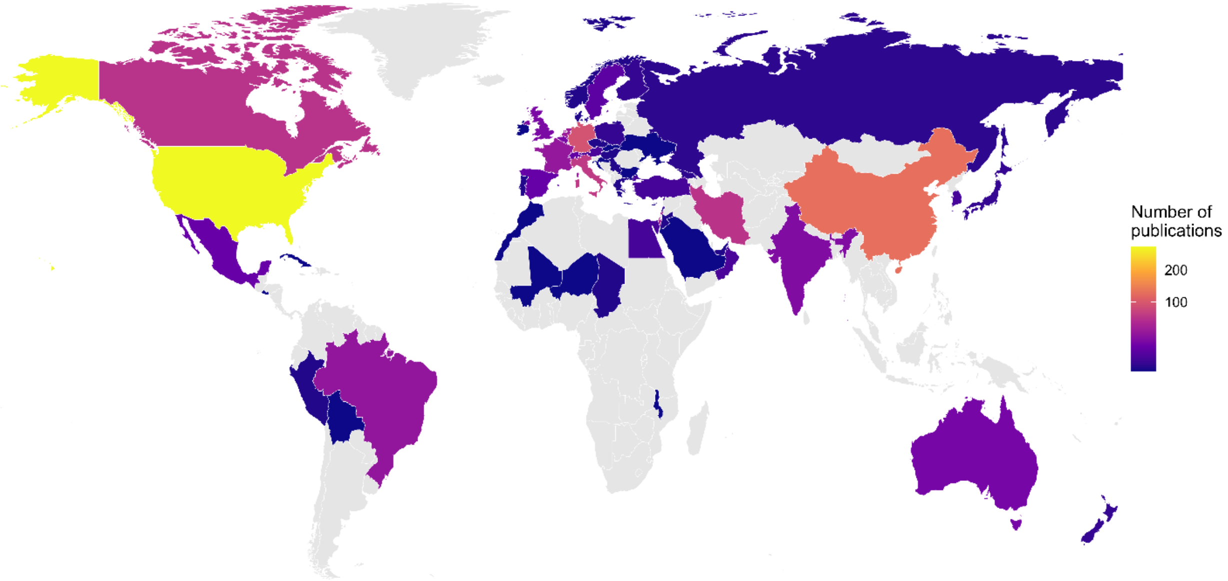

**Supplementary Figure S3.** Cumulative production of the top 6 journals.

Cumulative number of primary studies published in the six most productive journals (2000–2025). N = 810.

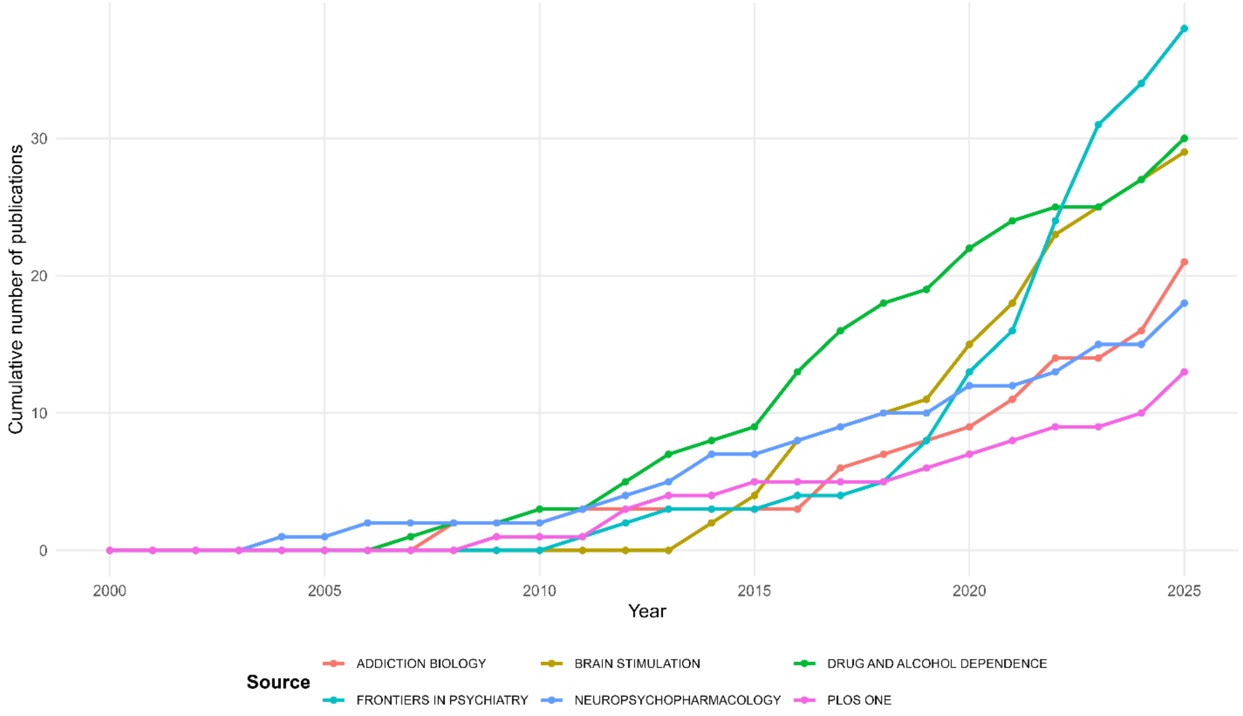

**Supplementary Figure S4.** Temporal trends in publications by substance category. Annual number of primary studies addressing each substance category (2000–2025). Categories are not mutually exclusive. N = 810.

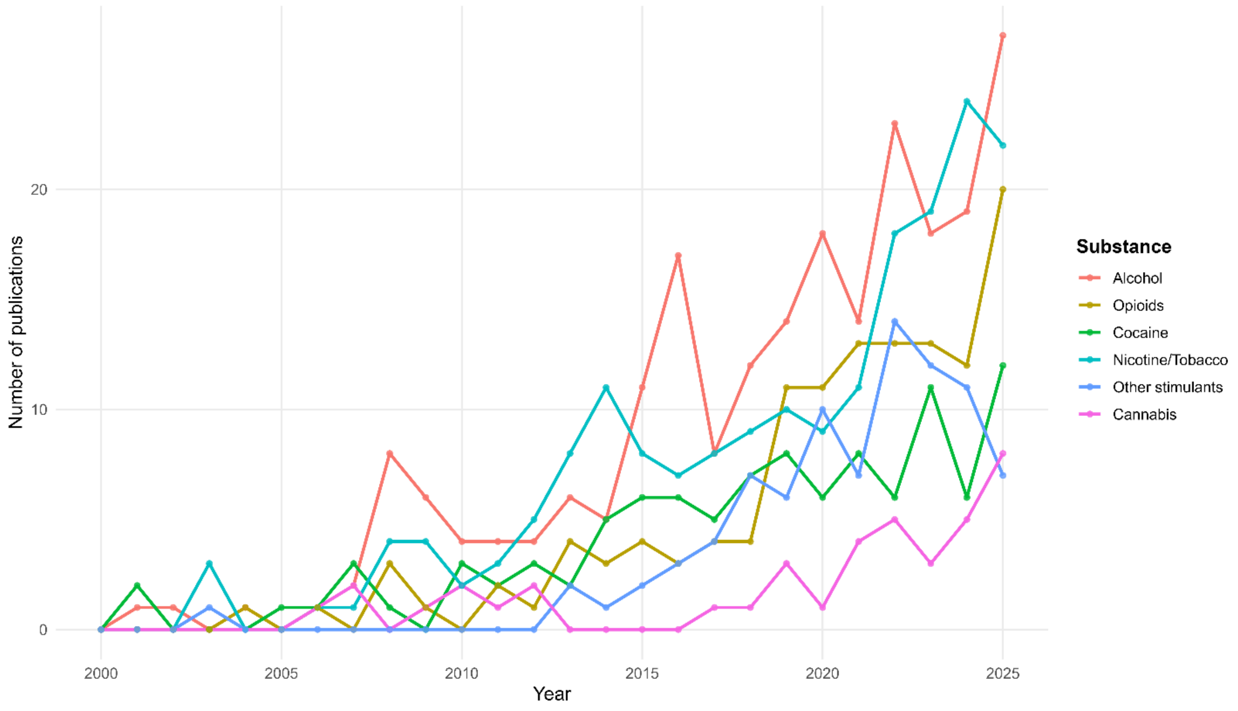

**Supplementary Figure S5.** Temporal trends in publications by neuromodulation modality. Annual number of primary studies for each neuromodulation modality (2000–2025). Modality categories are not mutually exclusive. N = 810.

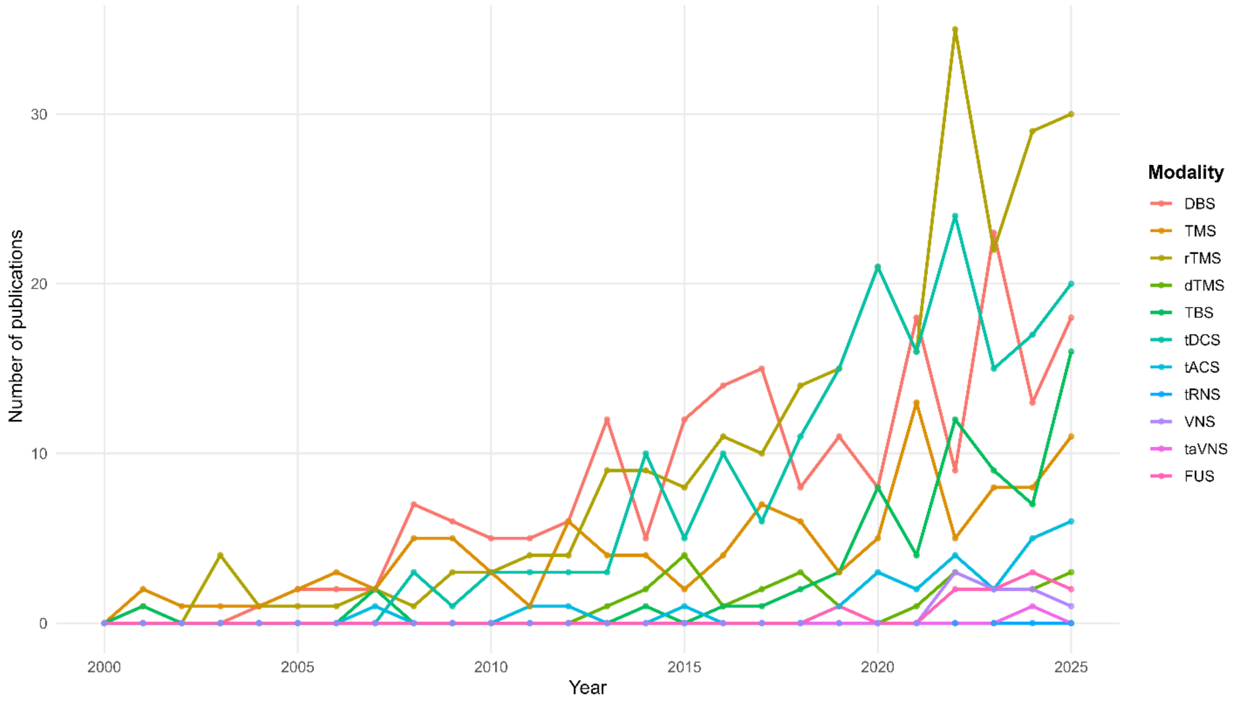

**Supplementary Figure S6.** Peak year of key author keywords.

Year of maximum annual frequency for each selected author keyword. Bubble size reflects peak frequency. In case of equal maxima, the most recent peak year is shown. N = 810.

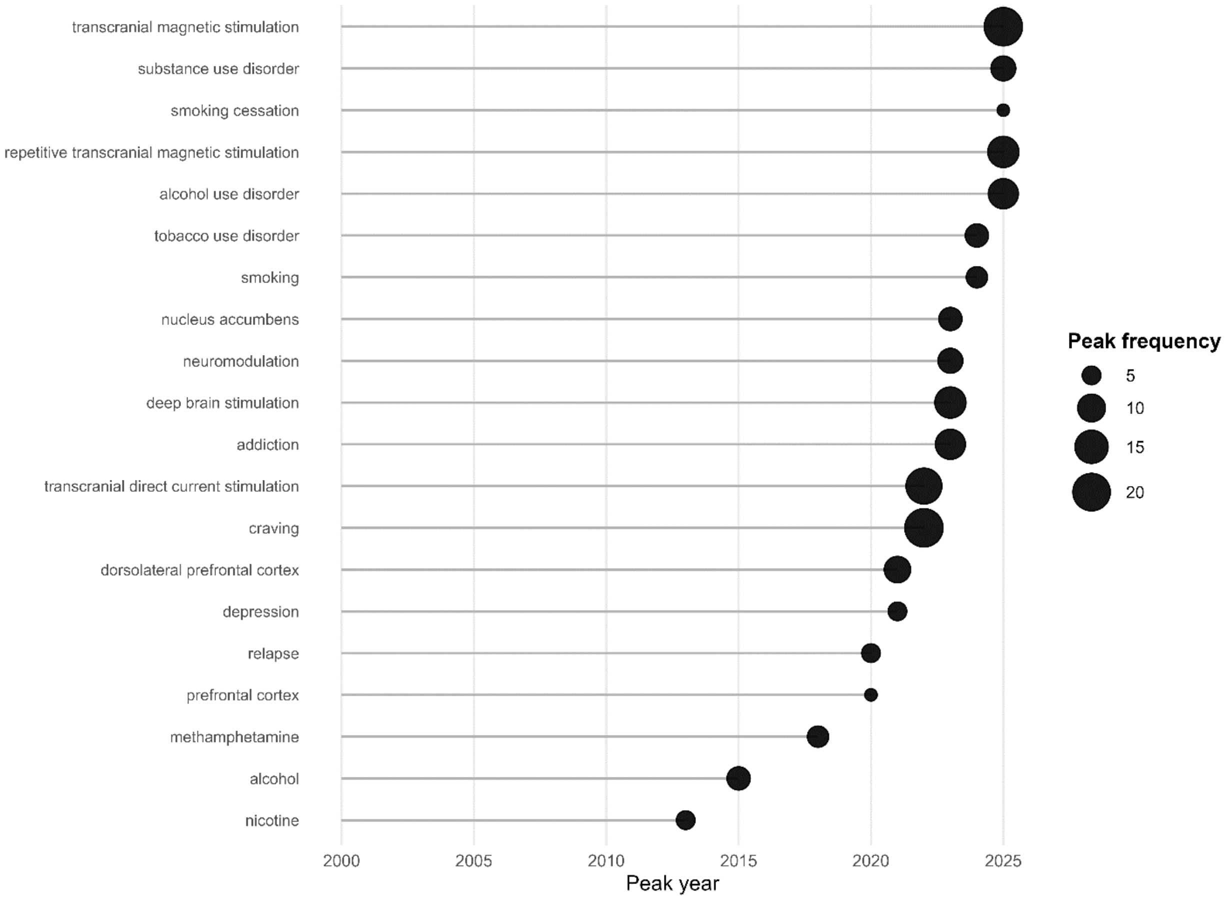
